# HECTOR: A Web-Based Tool for Automated *BRCA1/BRCA2* Variant Classification Under the ClinGen ENIGMA Specifications

**DOI:** 10.64898/2026.07.06.26357220

**Authors:** Tarık Düzenli, Ali Babazade, Ozan Vural, Yusuf Bahap, Mehmet Ali Ergün

## Abstract

**Background:** The ClinGen ENIGMA *BRCA1*/*BRCA2* Variant Curation Expert Panel (VCEP) has adapted the American College of Medical Genetics and Genomics/Association for Molecular Pathology (ACMG/AMP) framework into gene-specific specifications. However, applying these specifications manually remains labor-intensive and prone to inconsistency, requiring integration of population, computational, functional, and clinical evidence through gene-specific decision trees and a points-based classification system.

**Methods:** We developed HECTOR (HEreditary Cancer varianT Online Reclassifier), a free web-based tool that implements the complete ENIGMA VCEP v1.2 specifications for *BRCA1* and *BRCA2*. HECTOR automatically populates all evidence codes derivable from public data, routes curator-dependent evidence to a manual input layer and returns a transparent five-tier classification with code-level evidence. We validated HECTOR against two independent reference datasets: the 143-variant ENIGMA Evidence Repository, used as a clinical-grade reference standard, and 134 manually curated in-house variants of uncertain significance (132 unique variants). HECTOR was then applied to the complete ClinVar *BRCA1*/*BRCA2* catalog (n = 34,077).

**Results:** At the criterion level, HECTOR exactly reproduced 326 of 413 VCEP-assigned criteria (78.9%), with discordance arising predominantly from curator-dependent evidence rather than implementation errors. In an independent cohort of 134 manually curated in-house variants, HECTOR was fully concordant with expert consensus at the classification level and, even without auto-populated likelihood-ratio evidence, resolved more variants to a definitive classification than two generic ACMG/AMP classifiers. Across ClinVar, HECTOR classified 33,913 variants. Agreement with definitive ClinVar classifications was 96.7% for pathogenic variants and %72.7 for benign variants overall. Among variants for which HECTOR generated a definitive classification, directional concordance reached 99.7% for pathogenic and 99.9% for benign variants. HECTOR also resolved a substantial proportion of variants classified as uncertain (67.3%) or conflicting (88.7%), predominantly toward benign classifications.

**Conclusions:** HECTOR provides a faithful, transparent implementation of the ENIGMA VCEP v1.2 specifications for *BRCA1* and *BRCA2*, enabling rapid, standardized, and reproducible application of gene-specific variant classification guidelines while reducing the burden of manual curation.

## INTRODUCTION

Hereditary cancer syndromes caused by germline pathogenic variants account for approximately 10% of all cancers (Garutti et al., 2023). Germline pathogenic variants in *BRCA1* and *BRCA2* represent the most well-established genetic causes of hereditary breast and ovarian cancer syndrome. Pathogenic variants in *BRCA1* and *BRCA2* confer markedly elevated lifetime cancer risks. By age 80, the cumulative risk of breast cancer reaches 72% in *BRCA1* and 69% in *BRCA2* carriers, whereas the corresponding ovarian cancer risks are 44% and 17%, respectively (Kuchenbaecker et al., 2017). Accurate interpretation of *BRCA1/2* variants is therefore critical, as it directly influences risk-reducing surgical decisions, systemic therapy options such as PARP inhibitors, and cascade testing in at-risk relatives (Calabrese et al., 2024; O’Reilly et al., 2026).

The widespread implementation of next-generation sequencing has expanded access to multigene panel testing but has also led to a parallel increase in the detection of variants of uncertain significance (VUS) (Federici & Soddu, 2020; Rehm et al., 2023). According to the five-tier classification system proposed by the American College of Medical Genetics and Genomics/Association for Molecular Pathology (ACMG/AMP), VUS should not guide clinical decision-making (Richards et al., 2015). However, in real-world practice, uncertainty surrounding these variants may contribute to patient anxiety, inconsistent counseling, and, in some cases, inappropriate management strategies including unnecessary screening and surgeries (Neven et al., 2020; Rehm et al., 2023; Welsh et al., 2017). Importantly, VUS rates tend to be higher in underrepresented populations, reflecting limited population-specific data and highlighting disparities in variant interpretation (Dawood et al., 2024; Rehm et al., 2023). To address this, the ClinGen ENIGMA *BRCA1* and *BRCA2* Variant Curation Expert Panel (VCEP) developed gene-specific specifications that refine ACMG/AMP rules with statistically calibrated weights and explicit decision flowcharts (Parsons et al., 2024). Several studies have demonstrated substantial VUS reclassification, predominantly toward the benign spectrum when these specifications are applied (Benet-Pages et al., 2025; Innella et al., 2024; Kwong et al., 2026; Ozer et al., 2025).

Yet, the practical application of the ENIGMA VCEP specifications requires integrating data from multiple sources including specification tables, functional assay tables, multifactorial likelihood scores, population frequency databases, in silico prediction tools and reconciling them through gene-specific decision trees and a points-based combination framework. This multi-step workflow is labor-intensive and error-prone, particularly in routine clinical practice.

Existing tools partially address this. The UCSC Genome Browser *BRCA1*/*BRCA2* ENIGMA track set (Benet-Pages et al., 2025) consolidates many data layers in a single browser view but for limited number of variants and stops short of returning an integrated classification, leaving the analyst to combine the evidence manually. Although widely used, internet-based general-purpose variant classifiers apply the standard ACMG/AMP framework and do not faithfully reproduce the gene-specific modifications, and may therefore yield discordant classifications for the same variant. A dedicated tool that automates the entire workflow, from variant input to transparent, code-level classification, has not previously been available.

We here present HECTOR (HEreditary Cancer varianT Online Reclassifier), a user-friendly web-based *BRCA1*/*BRCA2* variant classifier implementing the complete ENIGMA VCEP V1.2 specifications. HECTOR automatically populates evidence codes derived from public data, including population frequencies, in silico predictions, functional assay results, and combined clinical likelihood ratios (LR), while reserving evidence that requires expert review, such as co-segregation and case-level data, for manual input. We validated HECTOR against two independent references, the 143-variant ENIGMA Evidence Repository as a clinical-grade gold standard, and 134 manually curated in-house VUS as a real-world consistency check. Additionally, we applied it across the full ClinVar *BRCA1*/*BRCA2* catalog (n = 34,077) to examine the current classification landscape under automated V1.2 application. HECTOR is freely available at https://hector.gazi.edu.tr.

## MATERIALS AND METHODS

### Engine Design and ENIGMA V1.2 Implementation

HECTOR implements the ClinGen ENIGMA *BRCA1* and *BRCA2* VCEP Classification Criteria V1.2, a gene-specific adaptation of the ACMG/AMP framework in which the applicability and weight of individual criteria are modified according to VCEP specifications. The tool is written in TypeScript and runs as a client–server web application. The user enters a variant in Human Genome Variation Society (HGVS) coding (c.) or protein (p.) notation, by genomic position, or as an exon-level deletion or duplication, and receives the evidence assigned at each criterion together with the resulting five-tier classification.

Automatically assigned evidence codes are grouped by data source. Population-frequency evidence is derived from the bundled non-cancer gnomAD datasets (v2.1.1 exomes and v3.1.2 genomes). BA1, BS1, and BS1_Supporting are assigned by comparing the FAF95 (filtering allele frequency) statistic, the 95% Poisson lower confidence bound of the population-specific allele frequency, with VCEP-defined thresholds, whereas PM2_Supporting is assigned when the variant is absent from these datasets. For the exome dataset, the published population-specific non-cancer FAF95 values are used directly. Because the genome dataset does not provide non-cancer FAF95 values, these were computed from the population-specific non-cancer allele counts using gnomAD’s filtering allele frequency algorithm (Hail’s filtering allele frequency). The highest FAF95 across both datasets and all assessed populations was then used for comparison with the VCEP-defined thresholds. The VCEP provision allowing larger insertions, deletions, and indels (>50 bp) to qualify for PM2_Supporting when absent from an appropriate dataset is not implemented because the specification does not specify which dataset should be used for variants larger than 50 bp.

Computational codes (PP3, BP4, BP7, and BP1_Strong) are derived from two bioinformatic predictors, BayesDel (Feng, 2017) (missense impact, applied within the clinically important domains) and SpliceAI (Jaganathan et al., 2019) (splicing impact) evaluated against the gene-specific thresholds and decision trees defined by the VCEP.

Predicted loss-of-function variants are evaluated using the PVS1 decision tree, with the assigned strength determined by nonsense-mediated decay competence, position relative to the final exon, and the potential for rescue transcripts. For premature termination variants, PM5 strength is similarly assigned on a per-exon basis according to the VCEP specification. At the obligate ±1,2 splice donor and splice acceptor dinucleotides, the PVS1 strength assigned by the decision tree is retained and is not downgraded on the basis of non-quantitative mRNA evidence (i.e., qualitative evidence of aberrant splicing without allele-specific quantification).

Functional assay evidence (PS3 and BS3) is assigned exclusively on the basis of functional data recommended by the ENIGMA VCEP. PS1 is evaluated by querying ClinVar, restricted to assertions from the ClinGen ENIGMA *BRCA1*/*BRCA2* VCEP, for a pathogenic variant with the same amino acid substitution (missense variants) or the same splice motif (splice variants). Legacy pre-VCEP ENIGMA classifications, which are based on International Agency for Research on Cancer (IARC) multifactorial posterior probabilities(Plon et al., 2008) rather than the ACMG/AMP framework, are identified by their submitting organization and are not used as PS1 references, consistent with the ENIGMA VCEP requirement that PS1 reference variants be classified under the ACMG/AMP framework. Nevertheless, HECTOR queries ClinVar for every variant and displays all available ClinVar classifications, irrespective of submitter, allowing classifications from non-VCEP sources to be considered during manual review.

Finally, a combined clinical likelihood ratio is calculated as the product of the case–control likelihood ratio (Zanti et al., 2025) and the ENIGMA literature-derived LR (Caputo et al., 2021; Easton et al., 2007; Li et al., 2020; Parsons et al., 2019) after correcting for evidence that would otherwise be counted in both terms. The case–control likelihood ratio is included only for variants assigned a calibrated evidence tier by the source; variants designated “No evidence”, indicating that the confidence interval is too wide to support any tier assignment, or “N/A” are excluded. For tier-assigned variants, the reported likelihood ratio is applied directly, without an additional significance filter based on the case–control odds ratio, consistent with ENIGMA V1.2 guidance recommending the case–control likelihood ratio in preference to the case–control odds ratio for PP4/BP5 assessment. The combined likelihood ratio is then mapped to the PP4 or BP5 evidence families according to the VCEP-defined thresholds, with values ≥2.08 supporting PP4-family codes and values ≤0.48 supporting BP5-family codes. PM3 is additionally assigned using the curated Fanconi anemia patient dataset provided in the specification, whereas the RNA-specific codes PVS1_RNA and BP7_Strong_RNA are assigned according to the VCEP splicing-evidence rubric.

The VCEP specification excludes a number of ACMG/AMP criteria from *BRCA1*/*BRCA2* classification, and these are therefore never applied: PS2, PM1, PM4, the original PM5, PM6, PP2, PP5, BP2, BP3, and BP6. Several additional criteria depend on evidence that cannot be assigned automatically. These include co-segregation data (PP1/BS4), traditional case–control odds ratios (PS4), biallelic phasing beyond the curated Fanconi anemia dataset, and a proband-specific phenotype likelihood ratio derived from the patient’s tumor pathology or clinical presentation rather than the population-level, literature-derived likelihood ratio incorporated automatically. Such case-specific evidence, including phenotype data, can be entered manually through a dedicated input layer integrated into the classification workflow.

Classification then proceeds using two parallel frameworks: the qualitative ACMG/AMP combining rules and the quantitative point-based system (Tavtigian et al., 2018; Tavtigian et al., 2020). Each applied criterion contributes points in the pathogenic or benign direction according to its assigned strength: ±1 for supporting, ±2 for moderate, ±4 for strong, and ±8 for very strong. The cumulative score is then mapped to the five-tier classification scale: pathogenic (≥ +10), likely pathogenic (+6 to +9), VUS (−1 to +5), likely benign (−6 to −2), and benign (≤ −7). When the ACMG/AMP framework yields a VUS but the point-based system yields a non-VUS classification, the point-based classification takes precedence, consistent with the VCEP recommendation that the quantitative framework be used to resolve otherwise conflicting evidence.

### Validation Tier 1 — ENIGMA Evidence Repository

The ENIGMA *BRCA1*/*BRCA2* VCEP maintains an Evidence Repository (https://erepo.genome.network) containing 143 *BRCA1*/*BRCA2* variants curated end-to-end under the V1.2 specification, with the complete evidence-code attribution for each variant published alongside the corresponding five-tier classification. Because these variants were curated under the same specification implemented by HECTOR, yet independently of it, the repository provides the closest available approximation to a clinical-grade reference standard for validating an automated classifier. We applied HECTOR to all 143 repository variants and compared its classifications with those of the VCEP at two levels: first, agreement in the five-tier classification (Pathogenic, Likely Pathogenic, VUS, Likely Benign, and Benign), and second, agreement in the individual evidence criteria assigned.

### Validation Tier 2 — In-House Manually Curated Cohort

To evaluate HECTOR under real-world diagnostic conditions, we retrospectively analyzed *BRCA1*/*BRCA2* variants identified in 1,614 patients who underwent hereditary cancer panel testing at the Gazi University Department of Medical Genetics between December 2022 and December 2023, all of whom met the National Comprehensive Cancer Network (NCCN) criteria for *BRCA1*/*2* testing. From this cohort, 134 variants that had originally been reported as VUS during routine clinical interpretation were selected and independently reclassified according to the ENIGMA *BRCA1*/*BRCA2* VCEP V1.2 specification by four experienced variant analysts. Discordant interpretations were resolved by consensus to establish a single reference classification for each variant. The same 134 variants were then analyzed using HECTOR, and the resulting classifications were compared with the manually curated reference at the five-tier level.

### Benchmarking against generic classifiers

For benchmarking, 132 unique in-house *BRCA1*/2 variants were evaluated using HECTOR under two conditions: with and without its auto-populated LR. The results were compared with those obtained from the generic variant interpretation platforms GeneBe (Stawinski & Ploski, 2024) and Franklin (Franklin by QIAGEN, 2026). Auto-populated LR comprised case-control likelihood ratios and ENIGMA literature-derived likelihood ratios retrieved automatically by HECTOR from publicly available resources; institution-specific tumor pathology LRs were not included, as these are not publicly accessible and are unavailable to both GeneBe and Franklin. Variant classifications from GeneBe and Franklin were retrieved through their public APIs on 22 April 2026 using the MANE Select transcripts (*BRCA1* NM_007294.4 and *BRCA2* NM_000059.4).

### ClinVar BRCA1/BRCA2 Full-Catalog Sweep

Beyond formal validation, we applied HECTOR to the complete ClinVar *BRCA1*/*BRCA2* catalog to characterize the current landscape of variant classification and assess its practical implications at scale. The complete ClinVar dataset for both genes was retrieved on 22 April 2026, yielding 15,839 *BRCA1* and 21,631 *BRCA2* records (37,470 in total), representing a snapshot of ClinVar at the time of retrieval. Before classification, 3,393 records were excluded, primarily because they lacked an aggregate clinical significance assignment, together with a smaller number whose HGVS descriptions could not be parsed by HECTOR (e.g., haplotype alleles and certain repeat notations). The remaining 34,077 variants were processed by HECTOR, of which 33,913 (99.5%) were successfully classified. The remaining 164 variants (0.5%) could not be resolved during HGVS parsing and therefore did not proceed to classification.

## RESULTS

### Evidence Repository

At the classification level, HECTOR agreed with the ENIGMA VCEP classification for 108 of the 143 variants (75.5%). Across these 143 *BRCA1*/*BRCA2* variants, the ENIGMA VCEP applied 413 evidence codes, of which HECTOR reproduced 326 (78.9%) at the identical code and strength (Figure 1). The remaining 87 discordant codes comprised 59 omitted by HECTOR and 28 assigned at a different strength. Of these, 78 (89.7%) represented curator-dependent evidence types that HECTOR does not assign automatically. Among the omitted codes, 25 reflected experimental mRNA or functional assay evidence (PVS1, PS3/BS3, and BP7), 21 reflected clinical or family-based evidence (PP4, PP1, BS2, and BP5), and four involved in-trans zygosity or Fanconi anemia phasing (PM3). All 28 strength discrepancies likewise arose from curator-dependent evidence, predominantly case–control likelihood ratios assigned one strength tier higher by HECTOR (PP4/BP5; 23 upgraded and 2 downgraded), together with two BP7 assignments up-weighted by the expert panel on the basis of mRNA evidence and one PVS1 assignment reduced from Very Strong to Strong pending confirmation of tandem orientation.

**Figure 1.**
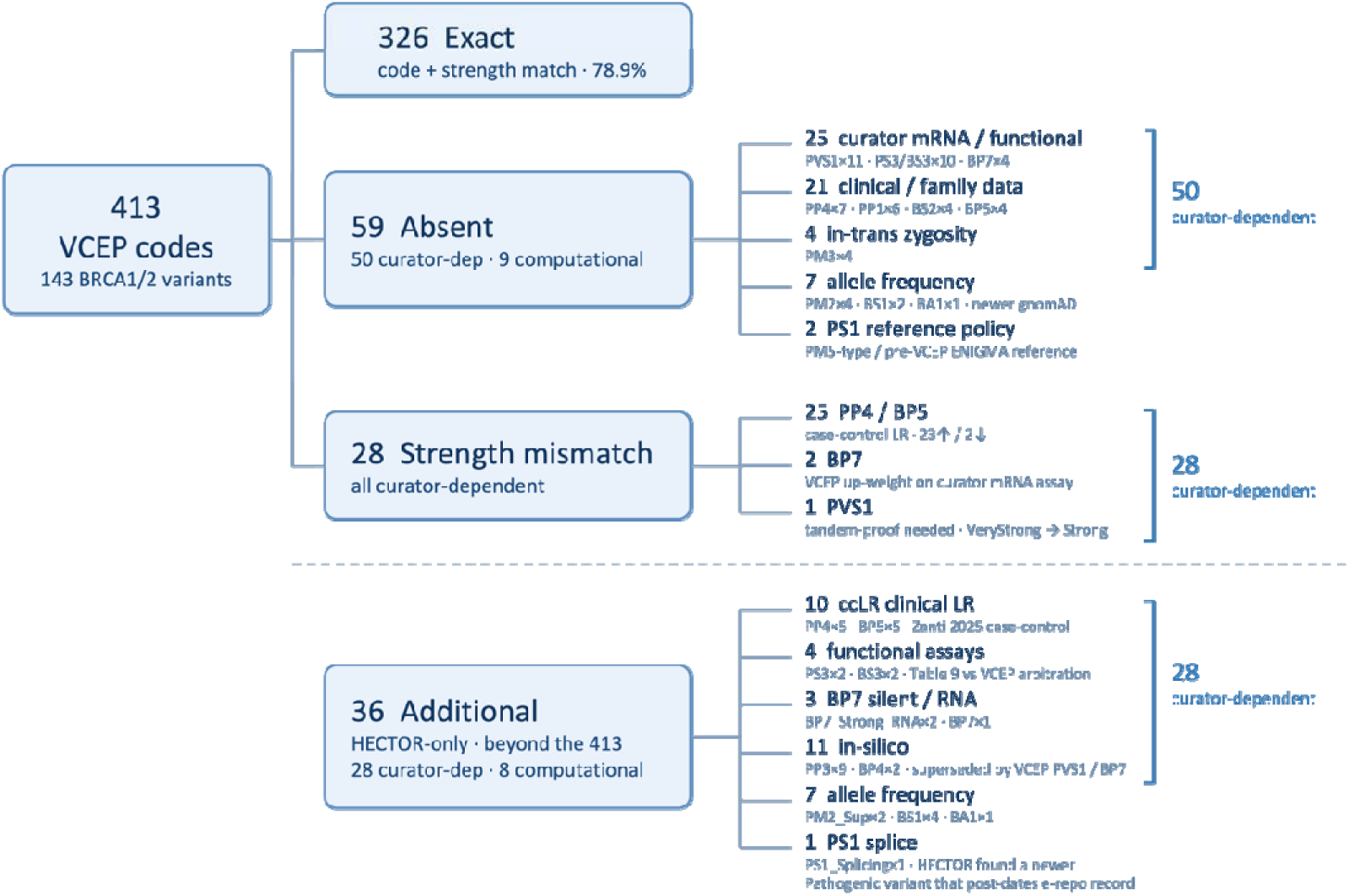
Concordance between HECTOR and the ENIGMA *BRCA1*/*BRCA2* Variant Curation Expert Panel (VCEP) on 143 variants from the ClinGen Evidence Repository.

The remaining nine discordant codes were computational. Seven were population-frequency codes (PM2 ×4, BS1 ×2, BA1 ×1) assigned by the VCEP using gnomAD v4.1. HECTOR is constrained by the ENIGMA V1.2 specification to gnomAD v2.1.1 (exomes) and v3.1.2 (genomes), and under these mandated datasets the variants did not meet the relevant frequency or absence thresholds. Accordingly, HECTOR’s assignments were specification-compliant, and the discordance reflects the panel’s occasional use of a newer population-frequency resource than that specified by ENIGMA V1.2.

The remaining two discordances reflected differences in PS1 reference availability and interpretation rather than incorrect application of the criterion. In one case (*BRCA1* c.5202T>G, p.Phe1734Leu), the VCEP assigned PS1 on the basis of a same-amino-acid reference variant (c.5200T>C, p.Phe1734Leu) that it had classified as Likely Pathogenic. Because this classification is not available in public ClinVar, where the variant remains a single-submitter VUS, HECTOR—which derives PS1 references exclusively from ClinVar—could not reproduce the assignment. In the other case (*BRCA1* c.4185G>C), the only potential reference variant (c.4185G>A) was represented solely by a pre-2020 ENIGMA submission based on the IARC multifactorial framework rather than ACMG/AMP classification. Because HECTOR restricts PS1 references to ACMG/AMP-classified ENIGMA VCEP submissions, the criterion was not assigned. Neither discordance represents an incorrect application of PS1; instead, both arise from differences in the evidence available to HECTOR and the reference policy applied.

Beyond the 413 evidence codes assigned by the VCEP, HECTOR applied an additional 36 codes that were absent from the repository annotations. These do not represent overcalling relative to the VCEP specification; in each case, the discrepancy can be traced to an identifiable source. Twenty-eight involved curator-dependent evidence derived from curated datasets that were either not cited by the VCEP or interpreted differently. These comprised five PP4 and five BP5 assignments based on the case–control likelihood ratio dataset, which HECTOR incorporates as an independent component of the combined clinical likelihood ratio; two PS3 and two BS3 assignments based on pre-curated functional studies that the VCEP interpreted differently in the presence of conflicting evidence; three RNA-related assignments (two BP7_Strong_RNA and one BP7); and eleven in silico prediction codes (PP3 ×9, BP4 ×2) retained only because HECTOR lacked the curator-derived RNA-based evidence (PVS1_RNA and BP7_Strong_RNA) that suppress underlying in silico codes.

The remaining eight additional codes were computational. Seven additional codes were population-frequency criteria (PM2_Supporting ×2, BS1 ×4, BA1 ×1) arising from three distinct sources. Two (PM2_Supporting) were the converse of the missing population-frequency codes, absent from HECTOR’s specification-mandated gnomAD v2.1.1/v3.1.2 but present in the panel’s v4.1. Two (one BA1, one BS1) were one-tier differences at the FAF95 threshold, where the panel applied the adjacent frequency tier rather than omitting the criterion. The remaining three BS1_Supporting codes fired on high-frequency alleles, two of them well-established pathogenic founder variants for which HECTOR applies the population-frequency code together with a founder-suspicion warning for curator review; none altered the final classification.

The final additional code was a single PS1_Splicing assignment. For *BRCA1* c.5152+3A>C, HECTOR identified a neighboring variant affecting the same splice-donor motif (c.5152+6T>C) that had subsequently been classified as Pathogenic in ClinVar. Because this ClinVar classification (August 2025) post-dated the frozen Evidence Repository record, the supporting PS1_Splicing evidence was unavailable to the panel during curation but was identified by HECTOR through real-time ClinVar querying. Detailed code-level comparisons for all variants are provided in the Supplementary Table S1.

### Inhouse

Across the 134 variant assessments (132 unique variants), encompassing 315 applied evidence codes, HECTOR and the manual analysts were fully concordant at the classification level and near-complete at the evidence-code level with only two discrepancies. Both arose from the same source: a third-party platform rounded BayesDel scores of 0.2989 and 0.2959 to 0.30, leading the curator to assign PP3, whereas HECTOR used the original unrounded values, which fall below the *BRCA2* threshold of 0.30, and therefore correctly did not assign the criterion.

Among the 132 unique variants, 107 (81.1%) had at least one available likelihood-ratio source, including case–control data, published multifactorial likelihood ratios, or institutional tumor pathology. HECTOR auto-populates the case–control and published multifactorial likelihood ratios and, where available, incorporated the institutional tumor-pathology likelihood ratio to generate a single combined clinical likelihood ratio. This crossed a VCEP evidence threshold for 64 of the 107 variants (59.8%), resulting in BP5 for 58 variants and PP4 for six, while the remaining 43 variants had likelihood ratios within the non-informative range (0.48–2.08) and therefore received neither code.

Incorporation of the combined clinical likelihood ratio changed the final classification of 32 of the 107 variants (29.9%), shifting 30 toward the benign spectrum and two toward the pathogenic spectrum. The benign-direction changes comprised 24 Likely Benign → Benign and six VUS → Likely Benign reclassifications. The two pathogenic-direction changes consisted of one Likely Benign → VUS and one Likely Benign → Likely Pathogenic reclassification (*BRCA1* c.5153-26A>G). Restricting the analysis to clinically meaningful boundary crossings—resolution of a VUS or a benign-to-pathogenic spectrum transition—the combined clinical likelihood ratio altered the classification of eight of the 107 variants (7.5%).

To benchmark HECTOR against generic variant interpretation platforms, classifications were compared with GeneBe and Franklin using the same set of 132 unique variants. With auto-populated likelihood-ratio evidence enabled, HECTOR remained fully concordant with expert manual curation, resolving 96 of 132 variants (72.7%) into definitive (non-VUS) classifications while leaving 36 as VUS (Table 1). In the absence of auto-populated LR evidence, HECTOR still classified 94 of 132 variants (71.2%) into definitive categories, compared with 85 of 132 (64.4%) by GeneBe and 31 of 132 (23.5%) by Franklin. Thus, even without LR evidence, HECTOR substantially outperformed both generic classifiers. Variant-level classification comparisons across HECTOR, manual curation, GeneBe, and Franklin are provided in Supplementary Table S2-S4.

**Table 1.** Five-tier classification of the 132 in-house *BRCA1*/*BRCA2* variants of uncertain significance by HECTOR, manual curation, GeneBe, and Franklin (QIAGEN). Each cell gives the number of variants assigned to the corresponding ACMG/AMP tier; the final row gives the number resolved to a definitive (non-VUS) classification. HECTOR is shown both with and without auto-populated likelihood-ratio (LR) evidence — the case–control and ENIGMA literature-derived LRs retrieved from public data; the institutional tumor-pathology LR is excluded throughout, as it is inaccessible to GeneBe and Franklin. Manual denotes the reconciled expert consensus of four analysts. GeneBe and Franklin were queried through their public APIs using the MANE Select transcripts (*BRCA1* NM_007294.4; *BRCA2* NM_000059.4). VUS: variant of uncertain significance

| Classification | Manual | HECTOR<br>(with LR) | HECTOR<br>(without LR) | GeneBe | Franklin |
| --- | --- | --- | --- | --- | --- |
| **Pathogenic** | 0 | 0 | 0 | 0 | 0 |
| **Likely pathogenic** | 1 | 1 | 0 | 1 | 2 |
| **VUS** | 36 | 36 | 38 | 47 | 101 |
| **Likely benign** | 60 | 60 | 82 | 80 | 25 |
| **Benign** | 35 | 35 | 12 | 4 | 4 |
| **Definitive (non-VUS)** | **96** | **96** | **94** | **85** | **31** |

### ClinVar

#### ClinVar Snapshot

At the time of retrieval, the ClinVar *BRCA1*/*BRCA2* catalog was broadly balanced between the two definitive classification directions (Figure 2). Of the 33,913 classified variants, 9,564 (28.2%) were classified as Pathogenic or Likely Pathogenic and 9,202 (27.1%) as Benign or Likely Benign, alongside 6,494 (19.1%) VUS, 8,580 (25.3%) variants with conflicting interpretations, and 73 (0.2%) assigned other classifications. The pathogenic spectrum comprised 8,377 Pathogenic, 607 Likely Pathogenic and 580 Pathogenic/Likely Pathogenic variants whereas the benign spectrum comprised 1,458 Benign, 7,546 Likely Benign and 198 Benign/Likely Benign variants.

**Figure 2.**
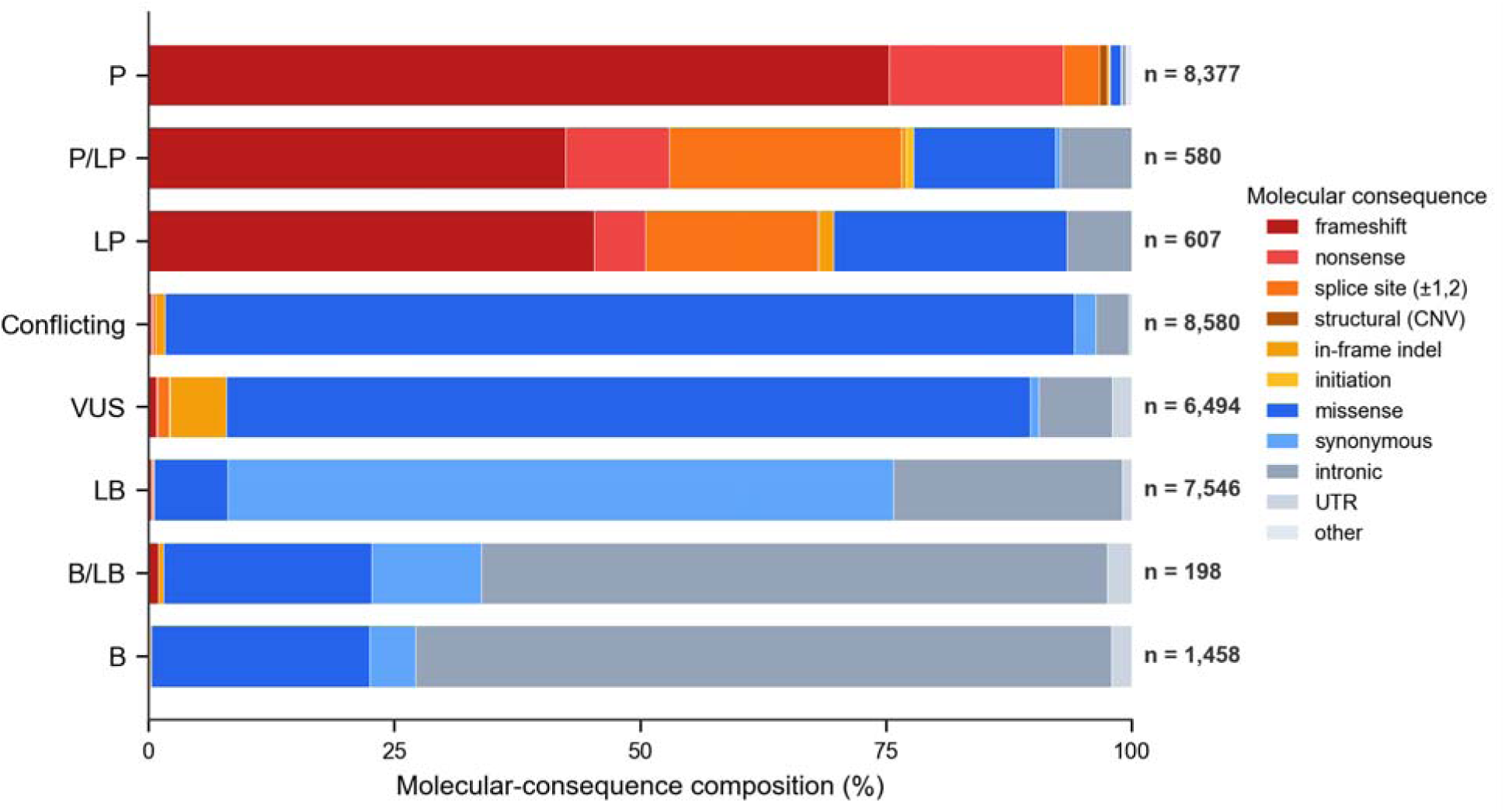
Molecular-consequence composition of the 33,840 classified ClinVar *BRCA1/BRCA2* variants (the 33,913 classified variants excluding 73 with a ClinVar ’other’ clinical significance — e.g., drug response or risk factor labels — that have no comparable five-tier assignment). Pathogenic and likely-pathogenic classes are dominated by protein-truncating consequences (frameshift, nonsense, splice site), whereas the VUS, Conflicting, and benign classes are dominated by missense and non-coding (synonymous, intronic) variants. (P, pathogenic; LP, likely pathogenic; VUS, variants of uncertain significance; LB, likely benign; B, benign).

Variant-type composition differed markedly across clinical-significance categories. Protein-truncating variants predominated among Pathogenic and Likely Pathogenic classifications, with frameshift (71.4%), nonsense (16.5%), and canonical splice-site variants (5.7%) collectively accounting for more than 90% of records, whereas missense variants represented only ∼3%. The pattern was reversed for VUS, with missense variants representing 81.7% of records, and became even more pronounced among variants with conflicting interpretations, where missense substitutions comprised 92.5% of records. In contrast, Benign and Likely Benign classifications consisted predominantly of synonymous (56%) and intronic (32%) variants, which together comprised nearly 90% of the cohort. Protein-truncating variants were virtually absent from this group. These classifications served as the baseline against which HECTOR reclassified the complete catalog under the ENIGMA VCEP V1.2 specification.

### Application across the *BRCA1*/*BRCA2* ClinVar catalog

Applied to the complete *BRCA1*/*BRCA2* ClinVar catalog, HECTOR generated a complete five-tier classification for 33,913 of 34,077 variants (99.5%). The remaining 164 variants (0.5%) were excluded at the input-parsing stage because their HGVS representations were not supported by the parser, primarily haplotype alleles and certain repeat notations, rather than because of failures during classification. The classified set comprised 12,939 *BRCA1* (38.2%) and 20,974 *BRCA2* (61.8%) variants, corresponding to a 1.6-fold excess of *BRCA2* variants.

HECTOR demonstrated substantially higher directional concordance with ClinVar for pathogenic than for benign classifications (Figure 3). Among the 9,564 variants classified by ClinVar as Pathogenic or Likely Pathogenic, 9,251 (96.7%) were assigned to the pathogeni spectrum by HECTOR. In comparison, 6,692 of 9,202 ClinVar Benign or Likely Benign variants (72.7%) were assigned to the benign spectrum. Restricting the analysis to variants for which HECTOR produced a non-VUS classification yielded near-complete agreement with ClinVar: 99.7% (9,251/9,275) in the pathogenic cohort and 99.9% (6,692/6,698) in the benign cohort. Thus, discordance with ClinVar arose almost exclusively from variants that HECTOR retained as VUS rather than from direct disagreement between pathogenic and benign classifications.

**Figure 3.**
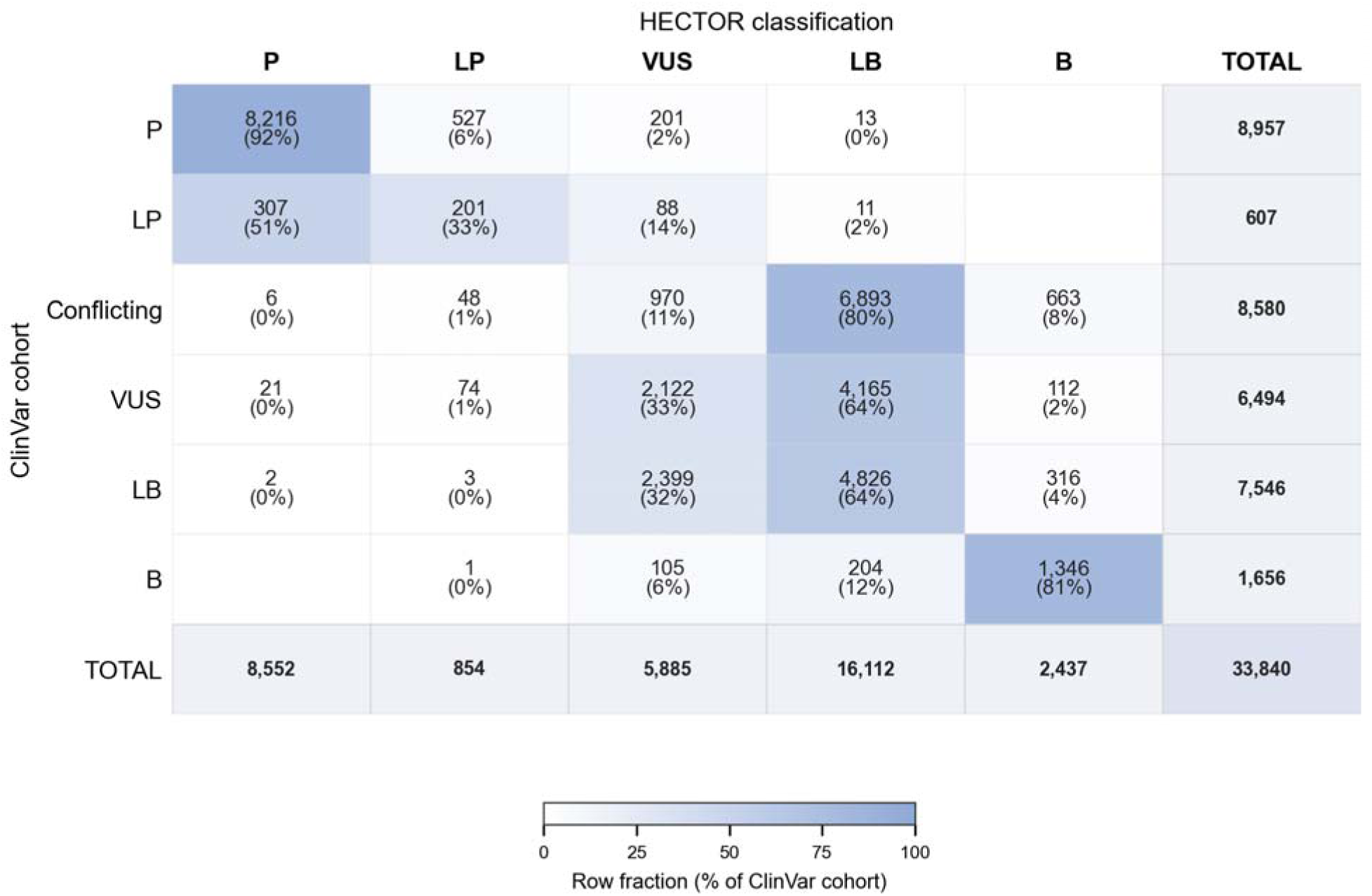
Confusion matrix of HECTOR classification (columns) against the ClinVar submission cohort (rows) for 33,840 *BRCA1*/*BRCA2* variants (the 33,913 classified variants excluding 73 with a ClinVar ’other’ clinical significance — e.g., drug response or risk factor labels — that have no comparable five-tier assignment). (P, pathogenic; LP, likely pathogenic; VUS, variants of uncertain significance; LB, likely benign; B, benign).

Across the full classified catalog, HECTOR reclassified variants with a marked bias toward the benign spectrum, with 11,857 benign-direction versus 155 pathogenic-direction assignments, representing an approximately 76-fold difference. Directional reversals were rare, comprising only 24 Pathogenic/Likely Pathogenic → Benign/Likely Benign and six Benign/Likely Benign → Pathogenic/Likely Pathogenic reclassifications. Most remaining discordant variants were conservatively retained as VUS, including 289 ClinVar Pathogenic/Likely Pathogenic variants and 2,504 ClinVar Benign/Likely Benign variants.

Among ClinVar Benign/Likely Benign variants that HECTOR retained as VUS, synonymous and intronic variants predominated, each accounting for 1,132 variants (45.2%), whereas missense variants represented only a small minority (160 variants, 6.4%). Among the synonymous variants, 1,001 (88.4%) were predicted to have no effect on splicing. Of these, all but one were located within HECTOR-defined functional domains and therefore were not eligible for BP1_Strong, instead receiving BP4 and BP7. Turning to the intronic variants, 1,057 (93.4%) were neither located at the canonical splice dinucleotides (+1, +2, −1, −2) nor predicted to affect splicing. Of these, 539 (51.0%) were deep intronic and 518 (49.0%) lay within the splice region. The distribution of these variants across the relevant VCEP decision trees is shown in Figure 4.

**Figure 4.**
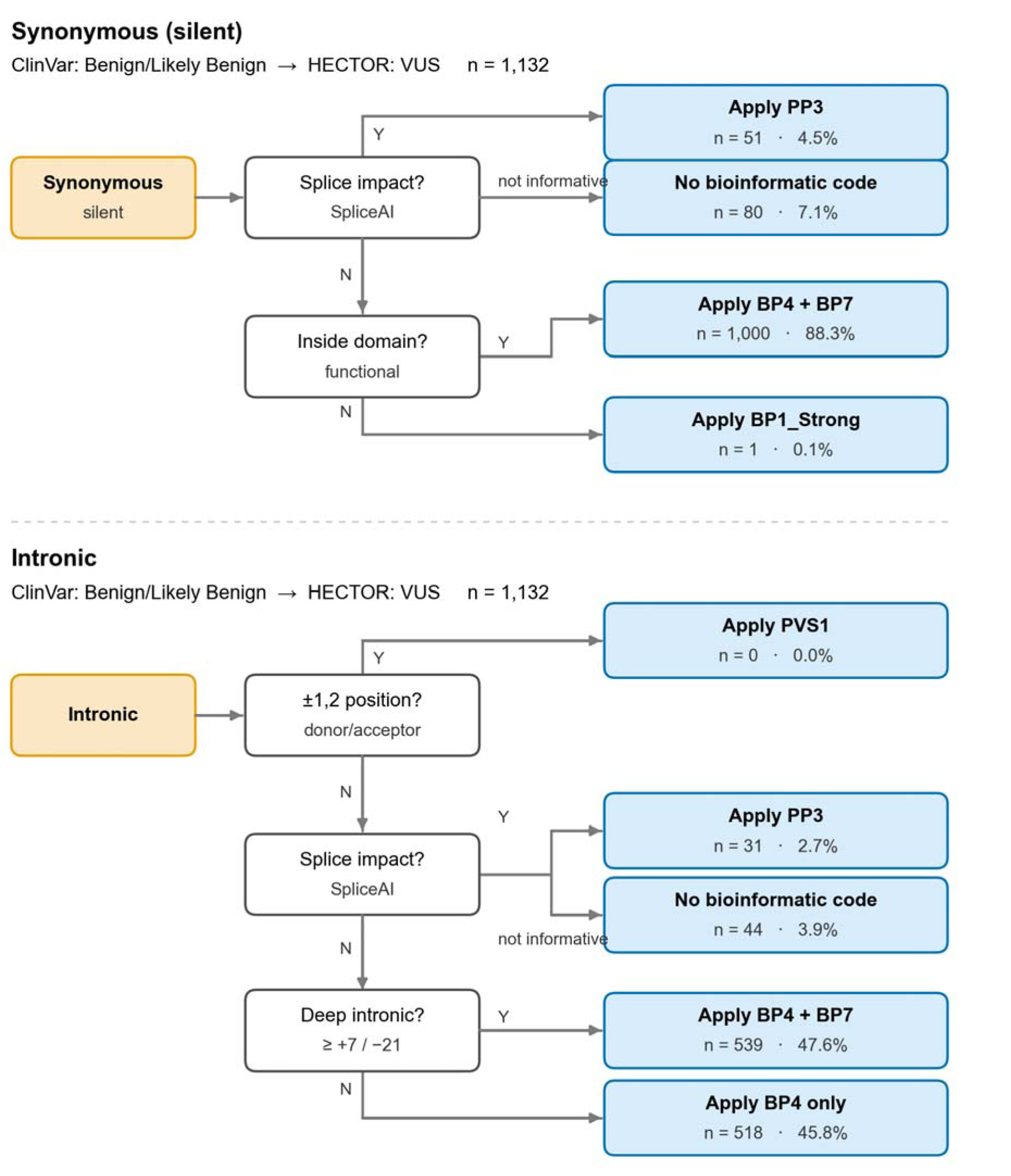
Bioinformatic decision path for the ClinVar Benign/Likely Benign *BRCA1*/*BRCA2* variants that HECTOR reclassified as VUS.

Because one of HECTOR’s pathogenic-direction criteria, PS1, relies on ClinVar—assigning evidence when a different nucleotide change producing the same amino acid substitution has already been classified as pathogenic in ClinVar—its concordance with ClinVar could, in principle, be partially self-reinforcing. To quantify this potential dependency, we reclassified all 9,251 pathogenic-direction variants after removing PS1. Only a single classification (<0.1%) changed, indicating that virtually all pathogenic-direction calls were supported by independent evidence. PM5_PTC, the same-residue premature-termination criterion, is likewise reference-based but derives from the ENIGMA VCEP specification rather than ClinVar. Removing both PS1 and PM5_PTC changed a further 16 classifications, none of which depended on ClinVar-derived evidence.

#### Resolution of uncertain and conflicting classifications

Beyond variants for which ClinVar already provides a definitive classification, a substantial proportion of the catalog remains clinically unresolved, comprising 6,494 variants classified as VUS and 8,580 variants with conflicting interpretations. When applied to these groups, HECTOR generated definitive (non-VUS) classifications for 67.3% (4,372/6,494) of VUS and 88.7% (7,610/8,580) of variants with conflicting interpretations (Figure 5). In both groups, these reclassifications were overwhelmingly toward benign interpretations. Among resolved VUS, 97.8% (4,277/4,372) were classified as likely benign or benign, whereas only 2.2% (95/4,372) were classified as likely pathogenic or pathogenic. A similar pattern was observed for variants with conflicting interpretations, among which 99.3% (7,556/7,610) were classified as likely benign or benign and only 0.7% (54/7,610) as likely pathogenic or pathogenic.

**Figure 5.**
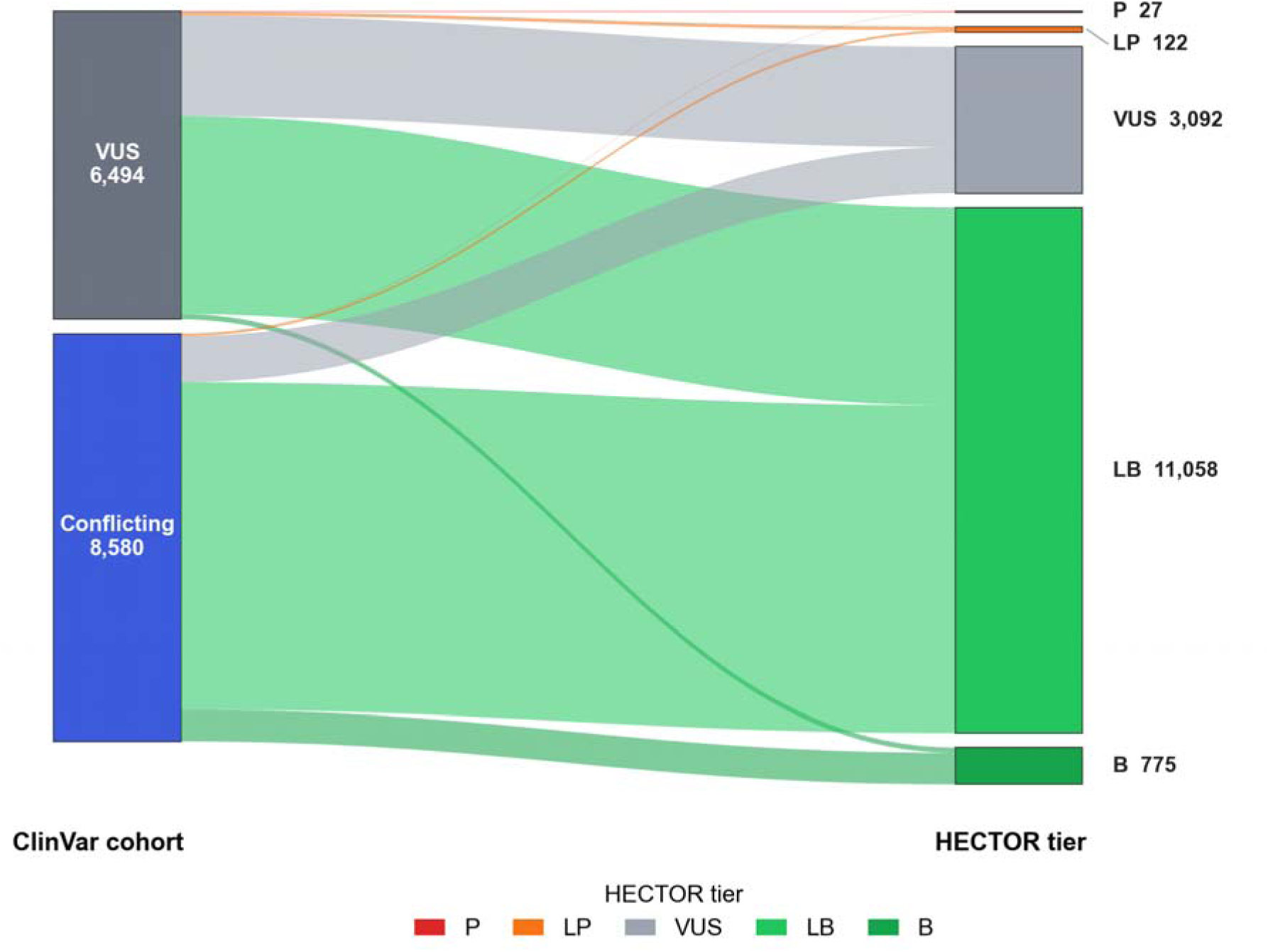
Reclassification of ClinVar VUS and Conflicting *BRCA1*/*BRCA2* variants by HECTOR. (P, pathogenic; LP, likely pathogenic; VUS, variants of uncertain significance; LB, likely benign; B, benign).

#### Molecular-consequence breakdown of the reclassified ClinVar VUS cohort

To investigate how HECTOR resolved variants of uncertain significance across different molecular consequence categories, we stratified the 6,494 ClinVar VUS by variant consequence (Figure 6). Missense variants accounted for the majority of the cohort (5,308/6,494, 81.7%), while the remaining 1,186 variants (18.3%) comprised non-missense consequences. Among missense variants, HECTOR reclassified 3,880 of 5,308 (73.1%), almost exclusively toward the benign spectrum: 3,843 (72.4%) were reclassified as likely benign or benign, whereas only 37 (0.7%) were upgraded to likely pathogenic or pathogenic; the remaining 1,428 (26.9%) retained a VUS classification. In contrast, 492 of 1,186 non-missense variants (41.5%) were reclassified, including 434 (36.6%) to likely benign or benign and 58 (4.9%) to likely pathogenic or pathogenic.

**Figure 6.**
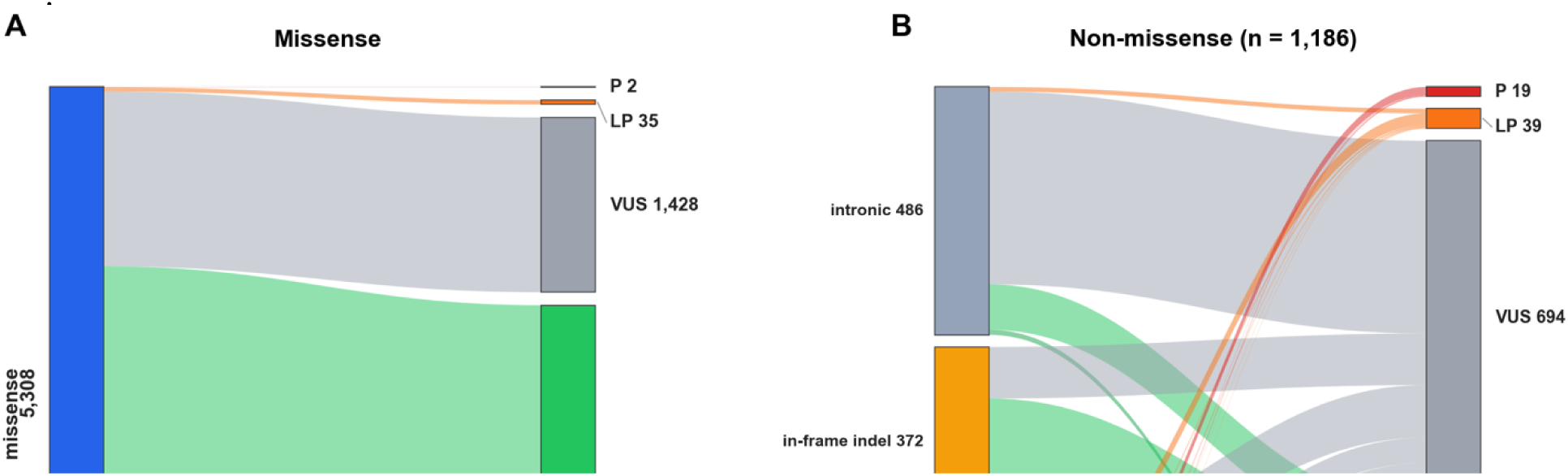
Molecular-consequence breakdown of the 6,494 ClinVar *BRCA1*/*BRCA2* variants of uncertain significance reclassified by HECTOR. Left panel, missense variants (n=5,308); right panel, non-missense variants (n=1,186), further partitioned by consequence class. Flows connect each consequence category (left) to the HECTOR five-tier classification (right) (P, pathogenic; LP, likely pathogenic; VUS, variant of uncertain significance; LB, likely benign; B, benign).

Within the non-missense group, intronic (n=486) and in-frame indel (n=372) variants were the most prevalent consequence types. Intronic variants proved the most resistant to reclassification, with 378 of 486 (77.8%) remaining VUS, whereas in-frame indels were resolved predominantly toward likely benign, with 269 of 372 (72.3%) receiving this classification. Synonymous variants (n=59) also tended to resolve toward likely benign (35/59, 59.3%), while untranslated region (UTR) variants (n=128) largely remained classified as VUS (103/128, 80.5%). Reclassifications toward pathogenicity were concentrated in variant classes with a higher prior likelihood of functional disruption, including frameshift variants (16 pathogenic and 2 likely pathogenic), splice-site variants (23 of 75 classified as likely pathogenic), nonsense variants (1 pathogenic and 1 likely pathogenic), as well as single initiation-codon and structural

#### Criterion-code application profile

Across the 33,913 classified variants, HECTOR applied ACMG/AMP criterion codes a total of 72,413 times, corresponding to an average of 2.14 criterion applications per variant (Figure 7). The most frequently applied was PM2_Supporting which was applied to 19,475 variants (57.4%). This was followed by BP1_Strong (15,178; 44.8%), PVS1 (9,144; 27.0%), the same-residue premature-termination criterion PM5_PTC (8,270; 24.4%)—which was tightly coupled with PVS1, co-firing in virtually all of its applications—BP4 (6,842; 20.2%), and BP7 (3,733; 11.0%).

**Figure 7.**
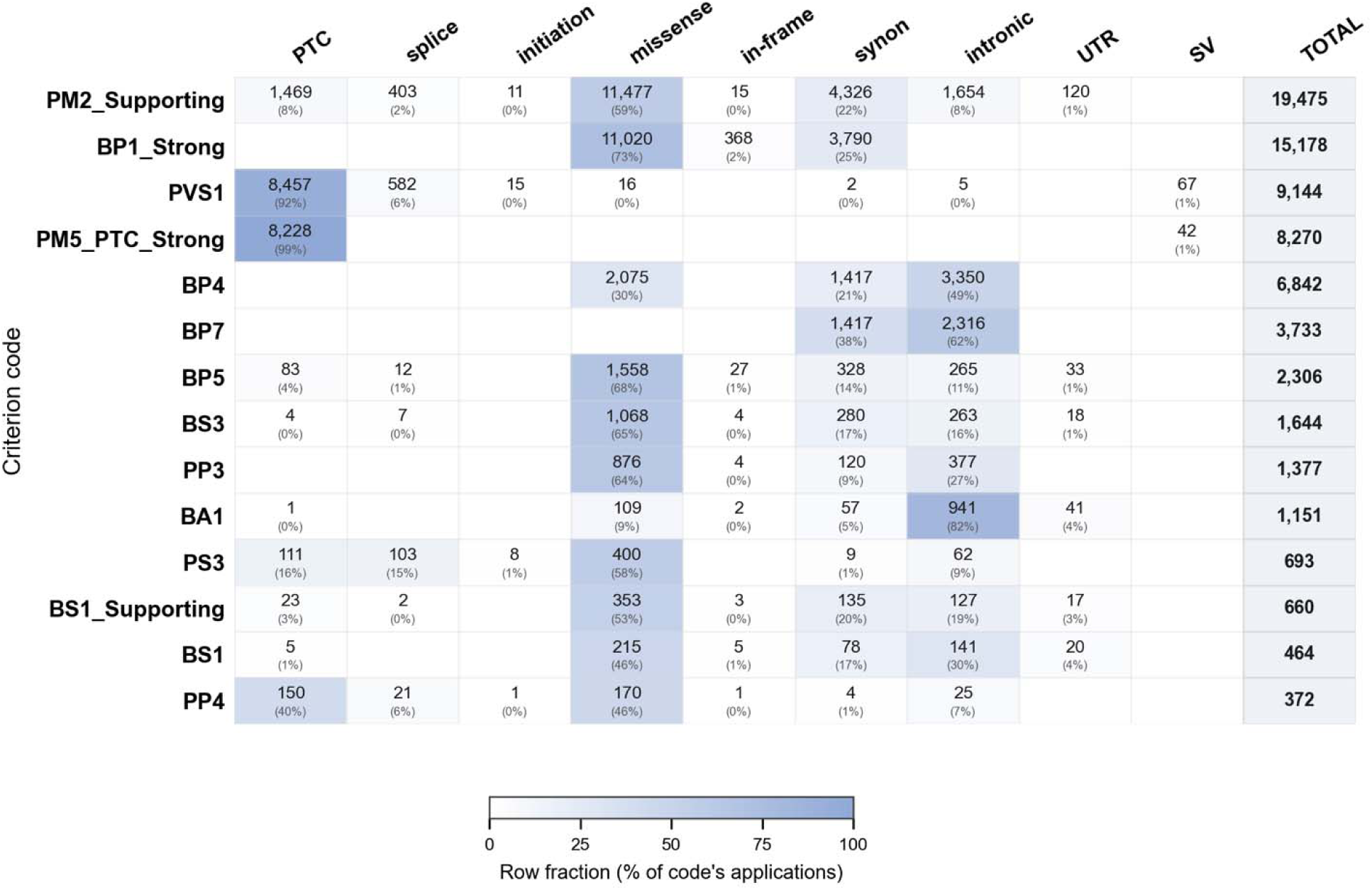
Application of the 14 most frequently used ENIGMA *BRCA1*/*BRCA2* VCEP V1.2 criterion codes (rows) across the 33,913 classified ClinVar variants, stratified by molecular consequence (columns).

Phenotype-derived evidence was available for 5,130 variants (15.1%), defined as those with at least one usable LR source. Among these, the combined LR reached the threshold for an ACMG/AMP criterion in 3,296 variants, triggering BP5 in 2,306 and PP4 in 990, while remaining neutral for the other 1,834 variants. Incorporation of LR evidence altered the final HECTOR classification of 1,064 of the 5,130 LR-bearing variants (20.7%), shifting 861 toward the benign direction and 203 toward the pathogenic direction. Most of these changes represented within-direction strengthening rather than reversals, including 648 Likely Benign-to-Benign and 95 Likely Pathogenic-to-Pathogenic transitions, whereas only nine variants were downgraded within the same direction (Pathogenic to Likely Pathogenic or Benign to Likely Benign). Of greater clinical relevance were the 312 changes that altered clinical actionability, among them 239 variants were resolved from VUS to a definitive classification (164 to Likely Benign, 29 to Benign, 27 to Likely Pathogenic, and 19 to Pathogenic), 71 previously definitive classifications were reclassified as VUS (57 Likely Benign, 12 Likely Pathogenic, and 2 Pathogenic), and two variants crossed the benign–pathogenic boundary (*BRCA1* c.5153-26A>G, likely benign to likely pathogenic and *BRCA1* c.5426T>A, likely pathogenic to likely benign).

## DISCUSSION

We developed HECTOR, an automated classifier that implements the ClinGen ENIGMA VCEP specification for *BRCA1* and *BRCA2* in full, populating every evidence criterion obtainable from public data and returning a five-tier call with its supporting codes and weights. We validated it against two independent references ENIGMA Evidence Repository and an in-house cohort. We then applied HECTOR to the full *BRCA1*/*BRCA2* ClinVar catalog to characterize the current state of these genes through a specification-faithful lens.

Against the ENIGMA Evidence Repository, HECTOR reproduced the expert panel’s exact classification tier for 75.5% of variants (108/143) and, at the evidence-code level, exactly reproduced 326 of 413 VCEP-assigned criteria (78.9%). Importantly, the remaining discordance reflected intrinsic limitations of automated evidence retrieval rather than differences in ACMG/AMP rule implementation. Code-level analysis showed that nearly all unrecovered criteria belonged to evidence categories inaccessible from public resources, including co-segregation data, phenotype likelihood ratios derived from individual cases, biallelic observations in non-public datasets, and functional or splicing assays published after the ENIGMA specification’s curation cutoff. Consequently, HECTOR recovered every VCEP criterion that was computationally accessible from the evidence permitted by the specification. We refer to this upper bound as the auto-population ceiling, representing the practical limit imposed by evidence availability rather than algorithmic performance. The few remaining discrepancies arose from evolving reference resources rather than missing curator knowledge or implementation errors; specifically, seven population-frequency criteria reflected VCEP evaluations based on newer gnomAD releases than the v2.1.1/v3.1.2 versions specified in the ENIGMA recommendations.

HECTOR’s concordance with ClinVar was markedly higher for pathogenic than for benign variants. The pathogenic-direction agreement reflects the architecture of pathogenicity in these genes: most pathogenic *BRCA1*/*BRCA2* variants are protein-truncating and thus fall within the scope of PVS1, a criterion HECTOR auto-populates directly, so the great majority reach a definitive pathogenic call without curator input. The benign-direction agreement was lower, driven almost entirely by variants ClinVar classifies as benign/likely benign but HECTOR conservatively retains as uncertain. On inspection, roughly nine in ten of these are intronic or synonymous changes and they present a particular classification challenge within the current framework. Most of the intronic variants are frequently absent from population databases (qualifying for PM2_Supporting), while SpliceAI often predicts no significant impact on splicing, resulting in BP4 and, for deep intronic variants, BP7 evidence which together total −1 or 0 points. Without additional lines of evidence which functional RNA-based evidence is rarely available, these variants cannot accumulate sufficient points to reach the −2 threshold required for likely benign classification under the ENIGMA specifications and remain VUS. This issue may indicate that benign in silico evidence for intronic variants remains underweighted in current practice. As demonstrated (Walker et al., 2023) variants with SpliceAI scores <0.1 showed a likelihood ratio of 0.059, corresponding to moderate benign evidence. Incorporation of such calibrated evidence strengths could improve classification yield for this variant category. The practical relevance of this limitation was also evident in our in-house reclassification cohort, where 26 of the 36 variants that remained classified as VUS were intronic, including 25 deep intronic variants, highlighting that this is not merely a theoretical limitation but a recurring challenge in routine variant interpretation.

These synonymous variants remaining in the VUS category typically receive PM2_Supporting due to their absence from population databases and are further supported by BP4 and BP7 evidence given their location within functional domains and lack of predicted impact on splicing, ultimately resulting in an overall VUS classification. We acknowledge that there is a small theoretical possibility that some of these variants may still affect translation kinetics through codon usage bias, potentially influencing co-translational folding within functionally important regions. However, examination of pathogenic/likely pathogenic synonymous variants in ClinVar (n = 7) indicates that all are located within splice regions, suggesting that disruption of splicing remains the predominant mechanism for clinically relevant synonymous changes. In this context, increasing the weight of SpliceAI-based evidence may be considered, particularly for better discrimination of truly splice-neutral variants.

The dominance of the two most frequently applied evidence codes reflects the composition of the *BRCA1*/*BRCA2* variant landscape itself. The widespread application of PM2_Supporting is driven by the fact that most catalogd variants are absent from reference population databases and therefore satisfy its population frequency criterion. Similarly, the prevalence of BP1_Strong is explained by the distribution of clinically important functional domains within the *BRCA1* and *BRCA2* proteins. Under the ENIGMA specifications, these domains encompass only 18.4% of *BRCA1* and 21.6% of *BRCA2*, leaving approximately four-fifths of each protein outside recognized functional domains. Consequently, the large proportion of missense variants occurring outside these domains falls within the scope of BP1_Strong.

Its prominence is further amplified by a gene-specific feature of the ENIGMA specification, which elevates the standard benign-supporting weight of BP1 to strong for *BRCA1*/*BRCA2*. This single code, contributing −4 points in the Tavtigian scoring system, is sufficient to classify a variant as likely benign even in the presence of PM2_Supporting (+1 point). The application of BP1_Strong to missense variants outside clinically relevant domains, resulting in their direct classification as likely benign, is therefore one of the most consequential features of the ENIGMA specifications. This reflects the well-established biological principle that disease-causing variants in *BRCA1* and *BRCA2* are predominantly protein-truncating and that missense variants outside defined functional domains have not been convincingly associated with increased cancer risk in clinical calibration studies. Consistent with this observation, 72 of the 76 pathogenic missense variants reported in ClinVar occur within ENIGMA-defined functional domains. The remaining four are located outside these domains but are all predicted to be pathogenic because of their effects on splicing rather than the resulting amino acid substitution. Thus, ClinVar currently provides no convincing evidence that amino acid substitution alone is sufficient to cause pathogenicity in missense variants outside ENIGMA-defined functional domains.

This code profile is the proximate explanation for the benign-leaning resolution of the uncertain and conflicting cohorts. Although the single most frequently applied code, PM2_Supporting, points toward pathogenicity, it carries only supporting weight; the benign-direction evidence firing across the out-of-domain missense pool — led by BP1_Strong at Strong weight and reinforced by BP4 and BP7 — is what actually carries those variants to a definitive benign call.

The integration of phenotype-related evidence through multifactorial likelihood analysis (PP4, BP5) represents one of the most distinctive features of the ENIGMA specifications. However, such evidence remains sparse. Automatically populated case-control and literature-derived LR evidence was available for only 15.1% of variants, but when present it changed the classification tier of approximately one in five variants (20.7%), predominantly toward the benign direction, and resolved 239 VUS to definitive classifications without curator input. In the in-house cohort, combined clinical LR evidence altered the final classification tier of only 32 of the 107 LR-bearing variants (29.9%), almost exclusively by reinforcing benign classifications and resolving VUS to likely benign, with only a single variant reclassified as likely benign to likely pathogenic. These findings suggest that expanding case-control, segregation, and phenotype datasets represents the most effective strategy for improving the yield of automated variant classification. In clinical practice, this evidence should therefore be actively collected and incorporated for each patient rather than relying on the limited information available in public databases.

Our implementation addresses one of the principal technical barriers to applying the ENIGMA specifications by automatically integrating phenotype-derived LR evidence from multiple heterogeneous sources into a unified interpretation framework. This removes the need for manual retrieval and harmonization of case-control, pathology, and other calibrated clinical evidence, substantially reducing the effort required for routine variant interpretation. Nevertheless, automation cannot compensate for the limited availability of the underlying data. For the majority of variants, particularly those that are rare or underrepresented in existing studies, calibrated phenotype-based LR evidence remains unavailable. Consequently, the primary limitation to broader implementation of phenotype-informed classification is no longer computational integration but the scarcity and incomplete population coverage of curated clinical datasets. Continued expansion and standardized sharing of phenotype-calibrated evidence across diverse populations will therefore be essential to fully realize the potential of automated ENIGMA-based variant classification.

Reassuringly, HECTOR’s advantage over generic classifiers does not depend on this evidence: even with all auto-populated likelihood-ratio evidence withheld, it resolved more in-house variants to a definitive classification than either GeneBe or Franklin, an advantage driven by faithful implementation of the gene-specific ENIGMA rules. Phenotype-derived evidence thus adds a further, complementary layer of resolution on top of an already robust rule-based foundation.

A number of limitations warrant consideration, most reflecting constraints of the underlying data and specification rather than the implementation itself. First, HECTOR auto-populates only the ACMG/AMP–ENIGMA evidence codes that can be derived from its bundled reference datasets and public APIs, while evidence requiring expert judgement, unpublished data, or case-/family-level information remains curator-dependent. Second, HECTOR calculates FAF95 using the same exact Poisson framework as gnomAD but without the per-sample read-depth ≥20 filter used by the ENIGMA VCEP, as this information is not available in the bundled gnomAD slice. Third, for whole-exon deletions and duplications, HECTOR currently applies only PVS1 and PM5_PTC because gnomAD-SV is not bundled, and more complex structural or rearrangement-level interpretations require manual review. Finally, because no comprehensive authoritative list of pathogenic founder alleles is provided in the specification, HECTOR does not suppress BS1 or BA1 for all established pathogenic founder variants, although such codes can be manually overridden by the curator.

In conclusion, HECTOR provides, to our knowledge, the first complete implementation of the ClinGen ENIGMA *BRCA1*/*BRCA2* VCEP v1.2 specifications, transforming a labor-intensive manual curation workflow into a transparent, code-level classification completed within seconds. Validation against the ENIGMA Evidence Repository demonstrated strong concordance, with the remaining discordance arising predominantly from curator-dependent evidence rather than implementation errors. Applied to the full ClinVar *BRCA1*/*BRCA2* catalog, HECTOR reproduced definitive classifications with high concordance and resolved a substantial proportion of variants with uncertain or conflicting interpretations, predominantly toward benign classifications. HECTOR is freely available at https://hector.gazi.edu.tr to facilitate rapid, transparent, and standardized expert variant interpretation.

## Data Availability

HECTOR is freely accessible at https://hector.gazi.edu.tr. The full per-variant output of the ClinVar BRCA1/BRCA2 sweep (vus-sweep-results.json), the e-repo validation bundle is explorable in hector.gazi.edu.tr.

## Acknowledgements

We would like to thank the Gazi University Directorate of Information Technologies, particularly Assoc. Prof. Dr. Ümit Atila, Head of the Department, and Information Technology Specialist Harun Keçeci, for their valuable correspondence and support. We also thank the patients for their participation and the ClinGen ENIGMA *BRCA1*/*BRCA2* Variant Curation Expert Panel for making their evidence repository publicly accessible.

## Notes

**Conflict of interest statement**, The authors declare no conflicts of interest.

### Competing Interest Statement

The authors have declared no competing interest.

### Author Declarations

Written informed consent was obtained from all participants in the in-house cohort. All analyses were performed in accordance with the Declaration of Helsinki. Ethical approval was obtained from the Gazi University Ethics Committee (03 March 2026, Decision No. 04; Research Code No. 2026-369). The ClinVar and ENIGMA Evidence Repository datasets used for validation and large-scale analyses are publicly available; no additional ethical approval was required for their use.

## REFERENCES

Benet-Pages, A., Laner, A., Nassar, L. R., Wohlfrom, T., Steinke-Lange, V., Haeussler, M., & Holinski-Feder, E. (2025). Reclassification of VUS in *BRCA1* and *BRCA2* using the new *BRCA1*/*BRCA2* ENIGMA track set demonstrates the superiority of ClinGen ENIGMA Expert Panel specifications over the standard ACMG/AMP classification system. Genet Med Open, 3, 101961. 10.1016/j.gimo.2024.101961

Calabrese, A., von Arx, C., Tafuti, A. A., Pensabene, M., & De Laurentiis, M. (2024). Prevention, diagnosis and clinical management of hereditary breast cancer beyond *BRCA1*/2 genes. Cancer Treat Rev, 129, 102785. 10.1016/j.ctrv.2024.102785

Caputo, S. M., Golmard, L., Leone, M., Damiola, F., Guillaud-Bataille, M., Revillion, F., Rouleau, E., Derive, N., Buisson, A., Basset, N., Schwartz, M., Vilquin, P., Garrec, C., Privat, M., Gay-Bellile, M., Abadie, C., Abidallah, K., Airaud, F., Allary, A. S., . . . Stoppa-Lyonnet, D. (2021). Classification of 101 *BRCA1* and *BRCA2* variants of uncertain significance by cosegregation study: A powerful approach. Am J Hum Genet, 108(10), 1907–1923. 10.1016/j.ajhg.2021.09.003

Dawood, M., Fayer, S., Pendyala, S., Post, M., Kalra, D., Patterson, K., Venner, E., Muffley, L. A., Fowler, D. M., Rubin, A. F., Posey, J. E., Plon, S. E., Lupski, J. R., Gibbs, R. A., Starita, L. M., Robles-Espinoza, C. D., Coyote-Maestas, W., & Gallego Romero, I. (2024). Using multiplexed functional data to reduce variant classification inequities in underrepresented populations. Genome Med, 16(1), 143. 10.1186/s13073-024-01392-7

Easton, D. F., Deffenbaugh, A. M., Pruss, D., Frye, C., Wenstrup, R. J., Allen-Brady, K., Tavtigian, S. V., Monteiro, A. N., Iversen, E. S., Couch, F. J., & Goldgar, D. E. (2007). A systematic genetic assessment of 1,433 sequence variants of unknown clinical significance in the *BRCA1* and *BRCA2* breast cancer-predisposition genes. Am J Hum Genet, 81(5), 873–883. 10.1086/521032

Federici, G., & Soddu, S. (2020). Variants of uncertain significance in the era of high-throughput genome sequencing: a lesson from breast and ovary cancers. J Exp Clin Cancer Res, 39(1), 46. 10.1186/s13046-020-01554-6

Feng, B. J. (2017). PERCH: A Unified Framework for Disease Gene Prioritization. Hum Mutat, 38(3), 243–251. 10.1002/humu.23158

Franklin by QIAGEN. (2026). https://franklin.genoox.com

Garutti, M., Foffano, L., Mazzeo, R., Michelotti, A., Da Ros, L., Viel, A., Miolo, G., Zambelli, A., & Puglisi, F. (2023). Hereditary Cancer Syndromes: A Comprehensive Review with a Visual Tool. Genes (Basel), 14(5). 10.3390/genes14051025

Innella, G., Ferrari, S., Miccoli, S., Luppi, E., Fortuno, C., Parsons, M. T., Spurdle, A. B., & Turchetti, D. (2024). Clinical implications of VUS reclassification in a single-centre series from application of ACMG/AMP classification rules specified for *BRCA1*/2. J Med Genet, 61(5), 483–489. 10.1136/jmg-2023-109694

Jaganathan, K., Kyriazopoulou Panagiotopoulou, S., McRae, J. F., Darbandi, S. F., Knowles, D., Li, Y. I., Kosmicki, J. A., Arbelaez, J., Cui, W., Schwartz, G. B., Chow, E. D., Kanterakis, E., Gao, H., Kia, A., Batzoglou, S., Sanders, S. J., & Farh, K. K. (2019). Predicting Splicing from Primary Sequence with Deep Learning. Cell, 176(3), 535–548 e524. 10.1016/j.cell.2018.12.015

Kuchenbaecker, K. B., Hopper, J. L., Barnes, D. R., Phillips, K. A., Mooij, T. M., Roos-Blom, M. J., Jervis, S., van Leeuwen, F. E., Milne, R. L., Andrieu, N., Goldgar, D. E., Terry, M. B., Rookus, M. A., Easton, D. F., Antoniou, A. C., Brca, Consortium, B. C., McGuffog, L., Evans, D. G., . . . Olsson, H. (2017). Risks of Breast, Ovarian, and Contralateral Breast Cancer for *BRCA1* and *BRCA2* Mutation Carriers. Jama, 317(23), 2402–2416. 10.1001/jama.2017.7112

Kwong, A., Ho, C. Y. S., Au, C. H., Ni, Z., Gan, Y., Tey, S. K., & Ma, E. S. K. (2026). Challenges of Real-World Utilization of Recommendations From ClinGen ENIGMA: A Focus on *BRCA2* Variant Classification in Chinese Population. JCO Precision Oncology, 10(1), e2500554. 10.1200/PO-25-00554

Li, H., LaDuca, H., Pesaran, T., Chao, E. C., Dolinsky, J. S., Parsons, M., Spurdle, A. B., Polley, E. C., Shimelis, H., Hart, S. N., Hu, C., Couch, F. J., & Goldgar, D. E. (2020). Classification of variants of uncertain significance in *BRCA1* and *BRCA2* using personal and family history of cancer from individuals in a large hereditary cancer multigene panel testing cohort. Genet Med, 22(4), 701–708. 10.1038/s41436-019-0729-1

Neven, P., Punie, K., Wildiers, H., Willers, N., Van Ongeval, C., Van Buggenhout, G., & Legius, E. (2020). Risk-reducing mastectomy in BRCA carriers: survival is not the issue. Breast Cancer Res Treat, 179(1), 251–252. 10.1007/s10549-019-05440-4

O’Reilly, C., McGarry, J. L., Zaborowski, A. M., Davey, M. G., Evoy, D., Rothwell, J., McCartan, D., Rutherford, C. L., Boland, M. R., & Prichard, R. S. (2026). Risk-Reducing Bilateral Mastectomy and Mortality in Carriers of *BRCA1* and *BRCA2* Variants: A Systematic Review and Meta-Analysis. JAMA Surg, 161(3), 260–267. 10.1001/jamasurg.2025.5929

Ozer, L., Aktuna, S., & Unsal, E. (2025). Reclassification of *BRCA1* and *BRCA2* Variants of Unknown Significance in a Turkish Cohort; A Single-Center, Retrospective Study. Eur J Breast Health, 21(4), 295–300. 10.4274/ejbh.galenos.2025.2025-5-2

Parsons, M. T., de la Hoya, M., Richardson, M. E., Tudini, E., Anderson, M., Berkofsky-Fessler, W., Caputo, S. M., Chan, R. C., Cline, M. S., Feng, B. J., Fortuno, C., Gomez-Garcia, E., Hadler, J., Hiraki, S., Holdren, M., Houdayer, C., Hruska, K., James, P., Karam, R., . . . Spurdle, A. B. (2024). Evidence-based recommendations for gene-specific ACMG/AMP variant classification from the ClinGen ENIGMA *BRCA1* and *BRCA2* Variant Curation Expert Panel. Am J Hum Genet, 111(9), 2044–2058. 10.1016/j.ajhg.2024.07.013

Parsons, M. T., Tudini, E., Li, H., Hahnen, E., Wappenschmidt, B., Feliubadalo, L., Aalfs, C. M., Agata, S., Aittomaki, K., Alducci, E., Alonso-Cerezo, M. C., Arnold, N., Auber, B., Austin, R., Azzollini, J., Balmana, J., Barbieri, E., Bartram, C. R., Blanco, A., . . . Spurdle, A. B. (2019). Large scale multifactorial likelihood quantitative analysis of *BRCA1* and *BRCA2* variants: An ENIGMA resource to support clinical variant classification. Hum Mutat, 40(9), 1557–1578. 10.1002/humu.23818

Plon, S. E., Eccles, D. M., Easton, D., Foulkes, W. D., Genuardi, M., Greenblatt, M. S., Hogervorst, F. B., Hoogerbrugge, N., Spurdle, A. B., Tavtigian, S. V., & Group, I. U. G. V. W. (2008). Sequence variant classification and reporting: recommendations for improving the interpretation of cancer susceptibility genetic test results. Hum Mutat, 29(11), 1282–1291. 10.1002/humu.20880

Rehm, H. L., Alaimo, J. T., Aradhya, S., Bayrak-Toydemir, P., Best, H., Brandon, R., Buchan, J. G., Chao, E. C., Chen, E., Clifford, J., Cohen, A. S. A., Conlin, L. K., Das, S., Davis, K. W., Del Gaudio, D., Del Viso, F., DiVincenzo, C., Eisenberg, M., Guidugli, L., . . . Medical Genome Initiative Steering, C. (2023). The landscape of reported VUS in multi-gene panel and genomic testing: Time for a change. Genet Med, 25(12), 100947. 10.1016/j.gim.2023.100947

Richards, S., Aziz, N., Bale, S., Bick, D., Das, S., Gastier-Foster, J., Grody, W. W., Hegde, M., Lyon, E., Spector, E., Voelkerding, K., Rehm, H. L., & Committee, A. L. Q. A. (2015). Standards and guidelines for the interpretation of sequence variants: a joint consensus recommendation of the American College of Medical Genetics and Genomics and the Association for Molecular Pathology. Genet Med, 17(5), 405–424. 10.1038/gim.2015.30

Stawinski, P., & Ploski, R. (2024). Genebe.net: Implementation and validation of an automatic ACMG variant pathogenicity criteria assignment. Clin Genet, 106(2), 119–126. 10.1111/cge.14516

Tavtigian, S. V., Greenblatt, M. S., Harrison, S. M., Nussbaum, R. L., Prabhu, S. A., Boucher, K. M., Biesecker, L. G., & ClinGen Sequence Variant Interpretation Working, G. (2018). Modeling the ACMG/AMP variant classification guidelines as a Bayesian classification framework. Genet Med, 20(9), 1054–1060. 10.1038/gim.2017.210

Tavtigian, S. V., Harrison, S. M., Boucher, K. M., & Biesecker, L. G. (2020). Fitting a naturally scaled point system to the ACMG/AMP variant classification guidelines. Hum Mutat, 41(10), 1734–1737. 10.1002/humu.24088

Walker, L. C., Hoya, M., Wiggins, G. A. R., Lindy, A., Vincent, L. M., Parsons, M. T., Canson, D. M., Bis-Brewer, D., Cass, A., Tchourbanov, A., Zimmermann, H., Byrne, A. B., Pesaran, T., Karam, R., Harrison, S. M., Spurdle, A. B., & ClinGen Sequence Variant Interpretation Working, G. (2023). Using the ACMG/AMP framework to capture evidence related to predicted and observed impact on splicing: Recommendations from the ClinGen SVI Splicing Subgroup. Am J Hum Genet, 110(7), 1046–1067. 10.1016/j.ajhg.2023.06.002

Welsh, J. L., Hoskin, T. L., Day, C. N., Thomas, A. S., Cogswell, J. A., Couch, F. J., & Boughey, J. C. (2017). Clinical Decision-Making in Patients with Variant of Uncertain Significance in *BRCA1* or *BRCA2* Genes. Ann Surg Oncol, 24(10), 3067–3072. 10.1245/s10434-017-5959-3

Zanti, M., O’Mahony, D. G., Parsons, M. T., Dorling, L., Dennis, J., Boddicker, N. J., Chen, W., Hu, C., Naven, M., Yiangou, K., Ahearn, T. U., Ambrosone, C. B., Andrulis, I. L., Antoniou, A. C., Auer, P. L., Baynes, C., Bodelon, C., Bogdanova, N. V., Bojesen, S. E., . . . kConFab, I. (2025). Analysis of more than 400,000 women provides case-control evidence for *BRCA1* and *BRCA2* variant classification. Nature Communications, 16(1), 4852. 10.1038/s41467-025-59979-6

